# A randomized, double-blind assessment of the effects of vortioxetine on functional connectivity in mild cognitive impairment

**DOI:** 10.64898/2025.12.10.25341891

**Authors:** M Klöbl, V Matev, P Stöhrmann, G Dörl, M Rothenberg, J Donath, C Cukaci, K Al Barede, R Lanzenberger, E Winkler-Pjrek, D Winkler

**Affiliations:** Department of Psychiatry and Psychotherapy, Medical University of Vienna, Vienna, Austria; Comprehensive Center for Clinical Neurosciences and Mental Health, Medical University of Vienna, Vienna, Austria; Karl Landsteiner University of Health Sciences, Dr. Karl-Dorrek-Straße 30, 3500, Krems, Austria; Division of Psychiatry and Psychotherapeutic Medicine, University Hospital Tulln, Alter Ziegelweg 10, 3430, Tulln, Austria

## Abstract

Vortioxetine has been reported to alleviate cognitive deficits in patients with depression, accompanied by changes in functional connectivity (FC). An open-label study suggested potential benefits of vortioxetine in patients with mild cognitive impairment (MCI), a condition also characterized by both cognitive and FC alterations. However, controlled studies are required to rule out nonspecific treatment effects and to determine whether FC changes are generalizable to this population.

To address this, we collected resting-state functional magnetic resonance imaging data from 47 patients with MCI at baseline, and after four and twelve weeks of randomized, double-blind treatment with either 10 mg or 20 mg vortioxetine, or placebo. Brain-wide FC changes were assessed using network-based R-statistics, an extension of the network-based statistics approach that allows mixed-effects modeling. Analyses were conducted with two different brain atlases to evaluate agreement across parcellations.

No vortioxetine-specific FC changes were observed. Instead, we identified a widespread network of predominantly reduced FC at four and twelve weeks across all groups (ps ≤ 0.012), with no additional significant changes between twelve and four weeks. Results showed high consistency across parcellations (rs ≥ 0.96).

These reductions in FC may reflect a reversal of preceding hyperconnectivity, potentially linked to compensatory network remodeling in MCI, as described in prior studies. While our findings do not support vortioxetine-specific FC effects in MCI, future research including healthy controls, pre-MCI assessments, and comparison groups with MCI and depression will be important to disentangle disease-related and treatment-related FC changes.

## Introduction

With a growing and aging global population, the incidence and prevalence of age-related disorders, accompanied by reductions in quality of life, are steadily increasing (Xi et al., 2025). This trend is expected to continue over the coming decades, with variations across geographic regions and socioeconomic strata (Xi et al., 2022). Mild cognitive impairment (MCI) is widely regarded as a prodromal stage of dementia and is characterized by memory impairment or cognitive deficits that exceed those expected in normal aging (Langa & Levine, 2014; Petersen et al., 1999). The well-established association between lower socioeconomic status and more severe age-related cognitive decline (Hahad et al., 2025; Wang et al., 2023) underscores the urgent need for accessible treatment strategies aimed at delaying, or ideally preventing, progression to dementia (Mitchell & Shiri-Feshki, 2009).

The antidepressant vortioxetine has been linked to cognitive improvements in patients with major depressive disorder across multiple studies (Katona et al., 2012; Mahableshwarkar et al., 2015; McIntyre et al., 2017; McIntyre et al., 2014). Notably, vortioxetine was the only antidepressant to demonstrate significant benefits in a meta-analysis of performance on the digit symbol substitution test in patients with depression (Baune et al., 2018). These findings have generated interest in vortioxetine as a potential disease-modifying treatment for MCI. Indeed, an open-label study in non-depressed patients with MCI reported promising improvements in several cognitive domains following vortioxetine treatment (Tan & Tan, 2021). However, the absence of a control group precludes definitive attribution of these effects to vortioxetine.

Structural and functional brain correlates of MCI have been extensively investigated using magnetic resonance imaging (MRI). Atrophy has been consistently observed in the hippocampus and other medial temporal lobe structures, as well as in the posterior cingulate, precuneus, and lateral temporal cortices (Pihlajamäki et al., 2009). More widespread cortical atrophy has also been reported in patients with MCI (Tabatabaei-Jafari et al., 2015). Functional MRI (fMRI) studies have revealed both increases and decreases in medial temporal lobe activation during the encoding of novel visual stimuli, findings that may reflect compensatory mechanisms in response to structural decline (Pihlajamäki et al., 2009). Abnormal functional connectivity (FC) has been particularly noted within the default mode network (DMN), especially in the posterior cingulate and precuneus (Pihlajamäki et al., 2009). DMN FC has therefore been extensively studied as a potential diagnostic marker distinguishing healthy individuals from those with MCI (Ibrahim et al., 2021). Vortioxetine has also been shown to modulate DMN activity in patients with major depressive disorder, reducing abnormally increased FC between the posterior cingulate and precuneus (Guo et al., 2025). In addition, vortioxetine attenuated activation during an n-back working memory task in the dorsolateral prefrontal cortex, hippocampus, insula, fusiform gyrus, and lingual gyrus in remitted patients with depression and healthy controls (Smith et al., 2018).

Building on these prior fMRI findings and addressing the lack of controlled studies on vortioxetine in MCI, we conducted a randomized, placebo-controlled trial over twelve weeks. We hypothesized that: (i) vortioxetine would induce differential changes in fMRI FC compared to placebo; (ii) early signs of FC adaptation would be detectable after four weeks and become more pronounced after twelve weeks; and (iii) a dose-response relationship would be observed, with greater FC changes at 20 mg compared to 10 mg vortioxetine.

## Methods

All procedures were conducted in accordance with the Declaration of Helsinki and the good scientific practice guidelines of the Medical University of Vienna (ethical committee number 1491/2019). The study was registered at the European Union Drug Regulating Authorities Clinical Trials Database (EudraCT) under the registration number 2019-001836-69.

### Study design

This was a randomized, double-blind, placebo-controlled trial investigating the effects of vortioxetine on MCI over 12 weeks. Potential participants underwent an initial screening, including routine blood tests, physical examination, and medical history assessment, including diagnosis of either hippocampal or medial temporal atrophy. Eligible subjects were randomized into three groups to receive either 10 mg vortioxetine, 20 mg vortioxetine, or placebo once daily. All participants assigned to vortioxetine began with 5 mg daily for three days. Those in the 20 mg group were then titrated to 10 mg for three days before reaching the final dose of 20 mg. Randomization was performed by a team member not involved in treatment administration. Blinding of the study medication was carried out by the Allerheiligen-Apotheke (Allerheiligenplatz 4, 1200 Wien). MRI data were acquired at baseline, after four weeks, and after twelve weeks of treatment initiation. Over the course of the study, MCI symptoms were repeatedly assessed via a neuropsychological test battery comprising the Alzheimer’s Disease Assessment Scale, the Rey Auditory Verbal Learning Test, the Digit Symbol Substitution Test, the Mini-Mental State Examination, the Clinical Global Impression of Severity, the Clinical Global Impression of Improvement, the CGI Efficacy Index, the Quality-of-Life Scale, and the Geriatric Depression Scale. The results of the neuropsychological tests will be published elsewhere.

### Sample size estimation

A meta-analysis reported the short-term antidepressant effect of vortioxetine compared with placebo with Cohen’s d = 0.217 (f = 0.11) (Pae et al., 2015). Cognitive improvements under vortioxetine relative to placebo were stronger, with z = [0.22, 0.52] (f = [0.22, 0.54]) (Harrison et al., 2016). This yields an average effect size of f = ((0.22 + 0.52) / 2 + 0.11) / 2 = 0.24. The predictive capabilities of FC for cognitive impairments related to Alzheimer’s disease was estimated with ρ = 0.49 (f = 0.56) (Lin et al., 2018). Averaging the effect sizes for vortioxetine and FC results in f = (0.24 + 0.56) / 2 = 0.4. Assuming 300 bilateral cortical neocortical regions (Van Essen et al., 2012), the alpha error of 0.05 was adjusted to α = 1.7 × 10^−4^ using the Sidak correction. Based on a repeated-measures ANOVA model with a within-between interaction effect in G*Power 3.1.9.2, three groups, three time points, a power of 1 - β = 0.80, and a temporal correlation of r = 0.5, the required sample size was N = 36. To allow for covariates and moderate deviations from assumptions, the target sample size was set at N = 45, corresponding to 15 participants per treatment group.

### Participants

Participants were recruited from the memory clinic of the Department of Psychiatry and Psychotherapy at the Medical University of Vienna and through advertisements in local newspapers. Inclusion criteria followed the National Institute on Aging–Alzheimer’s Association (NIA-AA) guidelines and comprised: age 50–80 years, subjective concern regarding cognitive changes, objective memory impairment relative to age group, and evidence of hippocampal or medial temporal atrophy. Additional requirements included adequate visual and auditory function for neuropsychological testing and the ability to provide written informed consent. Exclusion criteria were: current or past DSM 5 psychiatric disorder or severe neurological disorder, diagnosis of dementia, severe medical illness relevant to the study, use of prohibited medication, potential for childbearing, and contraindications to MRI. For each drop-out, another study participant was recruited.

### MRI acquisition

MRI data were acquired using a Siemens Prisma 3T scanner. Resting-state fMRI parameters were optimized according to Smith et al. (2013) as follows: TE / TR = 37 / 800 ms, 52° flip angle, multiband factor 8, 2 mm isotropic resolution, 104 x 104 voxel in-plane at 72 slices, resulting in a field of view of 208 x 208 x 144 mm, 2290 Hz/Px. Structural T1-weighted imaging was performed with the following parameters: TE / TR / TI = 2.22 / 2400 / 1000 ms, 8° flip angle, 0.8 mm isotropic resolution, 300 x 320 voxel in-plane at 208 slices, resulting in a field of view of 240 x 256 x 166.4 mm, 220 Hz/Px. Due to technical issues, one resting-state scan was acquired with an alternative sequence using a bandwidth of 2405 Hz/Px, while all other parameters remained identical.

### MRI processing

Resting-state fMRI data were preprocessed using the following steps: thermal noise reduction (Vizioli et al., 2021), slice-wise motion and physiological noise correction (Beall & Lowe, 2007, 2014), wavelet despiking (Patel et al., 2014), slice-timing correction, and distortion correction (Andersson et al., 2003). T1-weighted structural scans were processed using the longitudinal registration and segmentation pipeline in FreeSurfer 6. The longitudinal average of the structural scans was coregistered to the RS data. Parcellations based on the Desikan-Killiany-Tourville (DKT) atlas and the Destrieux atlas were applied to extract the first eigenvariate of the fMRI time series in subject space. This approach minimized variance introduced by normalizing brains with varying degrees of atrophy (Tabatabaei-Jafari et al., 2015). FC matrices were computed using partial correlations between all region-of-interest pairs, adjusted for white matter and cerebrospinal fluid signals via CompCor (Behzadi et al., 2007), and bandpass filtered to retain frequencies between 0.01 and 0.10 Hz (Hallquist et al., 2013). FC matrices were Fisher z-transformed to approximate normal distributions for statistical testing.

### Statistical testing

Given the known influence of parcellation choice on whole-brain FC analyses (Spies et al., 2019), all statistical tests were performed twice for validation, once using the DKT atlas and once using the Destrieux atlas.

### Model selection

Model selection was conducted to determine whether drug dose (placebo, 10 mg, 20 mg vortioxetine) and measurement time point (baseline, 4 weeks, 12 weeks) should be modeled as continuous variables or categorical factors. Continuous modeling provides parsimony and facilitates interpretation of dose-response and temporal effects, whereas categorical modeling allows for non-linear changes. To this end, the first principal component of the FC matrices was extracted and used as the dependent variable in the following mixed-effects model:

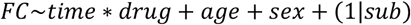

Here, *time* represented either discrete measurement points or continuous time, and *drug* denoted treatment group or dose (with placebo coded as 0 mg vortioxetine). *Age* and *sex* were included as covariates. A random intercept per *sub*ject was included, with further testing of a random time slope in case time was included as continuous variable. Candidate models were compared using likelihood ratio tests. Model construction and comparison were performed using the nlme package in R (Pinheiro et al., 2025). When likelihood ratio tests were non-significant, the more parsimonious model was selected.

### Network analysis

Functional network analysis was performed using Network-Based R-statistics (NBR; Gracia-Tabuenca and Alcauter (2020)), an extension of the classic network-based statistics approach (Zalesky et al., 2010). NBR employs linear mixed models to test effects across multiple connections, allowing inclusion of partially available datasets and simultaneous evaluation of both the size and overall strength of networks affected by fixed-effect experimental variables. Statistical inference was conducted via random permutation testing (1000 permutations) to achieve familywise error correction with additional Bonferroni adjustment for the number of comparison (three comparisons, each time point against each other) in case of significant results. We report p-values for both network size and strength.

The influence of individual regions of interest was quantified by averaging the absolute change of strength of all edges connected to each region within networks showing significant changes. To formally assess agreement across atlases, an FC score was calculated for each subject and session as the average across all edges with significant changes for each experimental variable. Partial correlations between these scores were then computed, adjusted for subject and session effects.

## Results

### Demographics

Including replacements for dropouts, 50 subjects were enrolled in the study. Of these, baseline MRI data was available for 47 participants, four-week MRI data for 41, twelve-week MRI data for 37 participants. Complete data was available from 34 subjects. The participants with available MRI data were 72.90 ± 10.49 years (median ± interquartile range) old and 13 identified as female. The neuropsychological tests showed quantitative improvements in all treatment groups (statistical results will be published separately). Demographic details are provided in Table 1.

**Table 1:** Participant demographics. IQR: interquartile range.

|  | Placebo | 10 mg vortioxetine | 20 mg vortioxetine | Overall |
| --- | --- | --- | --- | --- |
| Baseline MRI | 16 | 16 | 15 | 47 |
| Four-week MRI | 12 | 14 | 15 | 41 |
| Twelve-week MRI | 14 | 13 | 10 | 37 |
| Complete MRI data | 12 | 12 | 10 | 34 |
| Age [median ± IQR years] | 74.00 ± 7.45 | 72.80 ± 11.17 | 70.67 ± 10.18 | 72.90 ± 10.49 |
| Female | 4 | 5 | 7 | 13 |

### Network analysis

#### Model selection

Likelihood ratio tests showed no significant difference between modeling groups by continuous dose or as factor (DKT: p = 0.3126, Destrieux: p = 0.4784). However, model comparison strongly favored time as factor over a continuous variable (DKT: p = 0.0021, Destrieux: p = 0.0006). We thus modeled the treatment groups as continuous dose variable (0 mg (placebo), 10 mg, 20 mg vortioxetine) and time as factor (baseline, four weeks, twelve weeks).

#### NBR results

We found no significant influence of vortioxetine on FC over time (uncorrected ps ≥ 0.162). However, compared to baseline, there were strong FC changes across all groups four (ps ≤ 0.003) and twelve weeks (ps ≤ 0.012) into treatment, regardless of medication. There were no significant changes in FC between four and twelve weeks (ps ≥ 0.258). FC also strongly correlated with age (ps < 0.003). The significant results showed excellent agreement across both atlases (rs ≥ 0.96). The p-values for all variables are presented in Table 2.

**Table 2:** Network-Based R-statistics results. p-values for the Desikan-Killiany-Tourville (DKT) and Destrieux atlas are provided together with partial correlations of the averaged functional connectivities per subject and session for each significant network. Multiple p-values for the same atlas and same variable mean that separate disjunct networks were identified.

|  | DKT |  | Destrieux |  | Agreement [r] |
| --- | --- | --- | --- | --- | --- |
|  | Number of components [p] | Network strength [p] | Number of components [p] | Network strength [p] |  |
| Dose | 0.876 | 0.875 | 0.564 | 0.554 |  |
| Four weeks vs. baseline | 0.003 (0.001) | < 0.003 (0.001) | < 0.003 (0.001) | < 0.003 (0.001) | 0.98 |
| Twelve weeks vs. baseline | 0.006 (0.002) | 0.003 (0.001) | 0.012 (0.004) | < 0.003 (0.001) | 0.98 |
| Twelve weeks vs. four weeks | 0.258 | 0.288 | 0.315 | 0.294 |  |
| Age | < 0.003 (0.001) | < 0.003 (0.001) | < 0.003 (0.001) | < 0.003 (0.001) | 0.96 |
| Sex | 0.432 | 0.297 |  |  |  |
|  | > 0.999 | > 0.999 | 0.107 | 0.129 |  |
| Dose at four weeks vs. baseline | 0.162 | 0.230 | 0.205 | 0.209 |  |
| Dose at twelve weeks vs. baseline | 0.649 | 0.608 |  |  |  |
|  | > 0.999 | > 0.999 | 0.524 | 0.564 |  |
| Dose at twelve weeks vs. four weeks | 0.180 | 0.188 | 0.228 | 0.230 |  |

Four weeks and, slightly less so, after twelve weeks after treatment initiation the functional connectivity of a large network throughout the brain mainly showed a marked decrease across all treatment groups (placebo, 10 mg, or 20 mg vortioxetine). The networks additionally contained some edges with increasing FC over time. The same pattern of mainly decreasing FC was observed with higher age. The results are depicted in Figure 1 for the DKT atlas and the average network changes in Figure 2.

**Figure 1:**
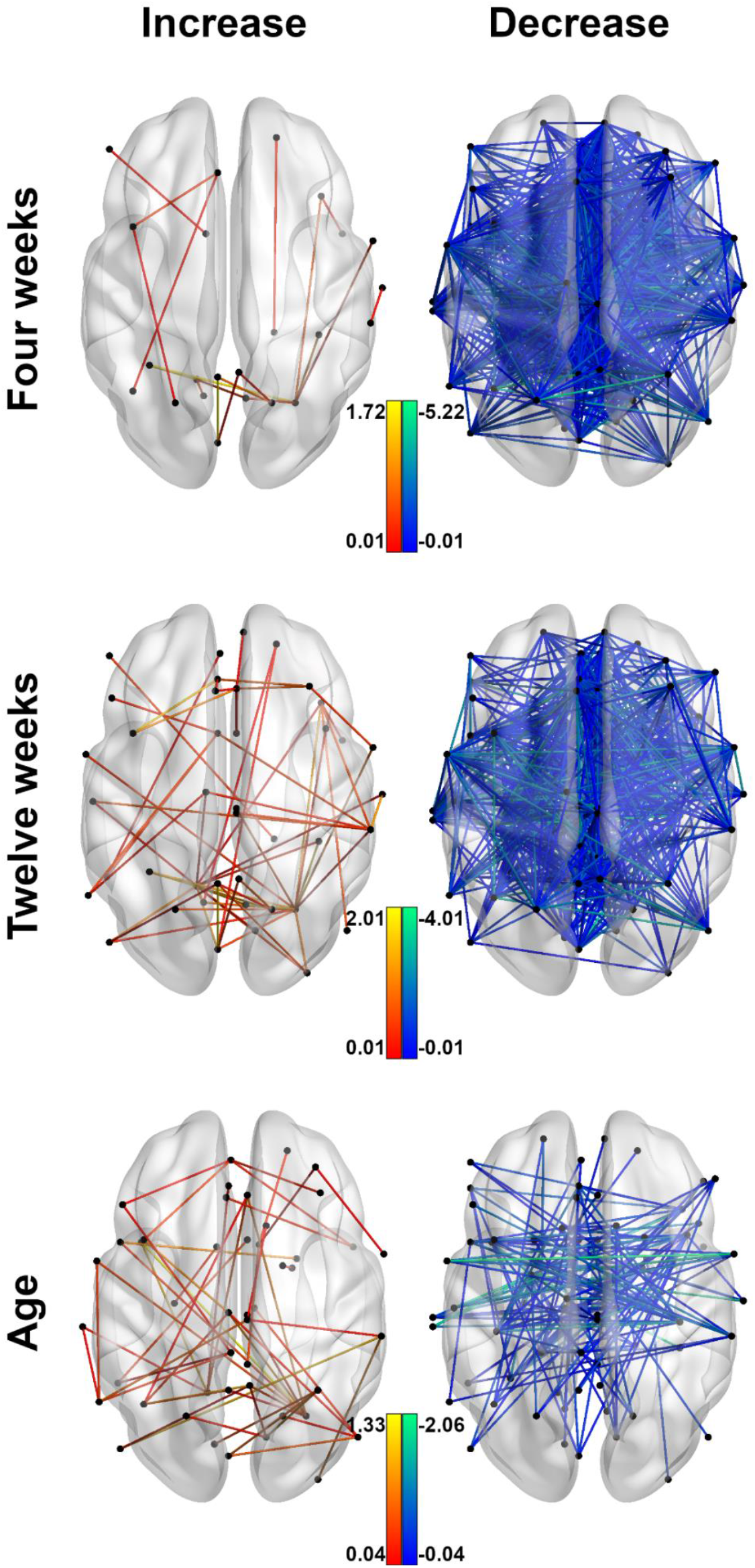
Significant results of the Network-Based R-statistics analysis for the Desikan-Killiany-Tourville atlas.The graphs show the edges of the networks of functional connectivity changes four and twelve weeks after treatment initiation and the network correlating with participant age. For better readability, the positive and negative edges of the same networks are presented separately.

**Figure 2:**
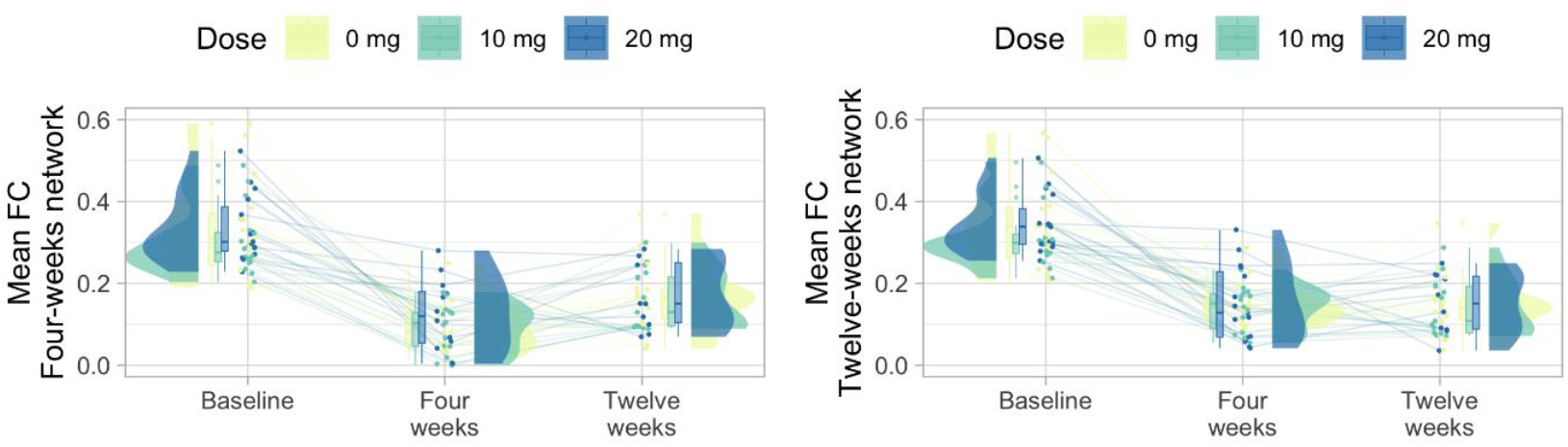
Time course of average functional connectivity (FC) for networks with significant changes.Each point represents the average FC per subject across the network with significant changes after four (left) and twelve weeks (right). While the networks contained some edges with increasing FC over time, the overall reduction in FC from baseline to for weeks into treatment is clearly visible.

#### Most influential nodes

For the DKT atlas, the strongest increase in connected edges was observed for the FC of the right cerebellum between baseline and four as well as twelve weeks into treatment. The left cerebellum showed similar and only slightly weaker effects. The FC of the right cerebellum also showed the strongest increase with age.

The top 10% nodes with the decrease in strength of connected edges for the DKT atlas were as follows: The left precentral gyrus as well as the left and right transverse temporal gyri showed strong changes after four and twelve weeks, but also in relation to age. Furthermore, the right posterior cingulate cortex was among the top nodes with FC changes after four and twelve weeks. The left postcentral gyrus and right superior temporal gyrus were implicated after four weeks and strongly correlated with age. Connections from the left posterior cingulate cortex and right insula showed marked FC decreases after twelve weeks. Furthermore, after twelve weeks of treatment, the left caudal anterior and isthmus cingulate as well as the left insula and right precentral gyrus showed strong decreases in FC. Lastly, the left entorhinal cortex, fusiform gyrus, and left amygdala showed a strong negative correlation with age. The results for the Destrieux were similar but distributed over more regions due to the more fine-grained parcellation.

## Discussion

Vortioxetine has been reported to improve cognitive deficits in patients with depression and in non-depressed individuals with MCI. Both MCI and vortioxetine treatment have been associated with alterations in brain FC. However, no placebo-controlled study on the effects of vortioxetine in patients with MCI has been conducted so far. Moreover, until now, no placebo-controlled study has examined vortioxetine’s effects on FC in patients with MCI. Contrary to our initial hypotheses, we did not observe significant differences in FC changes between patients receiving vortioxetine (10 or 20 mg) and those receiving placebo. Instead, we identified a widespread network of predominantly reduced FC across all treatment groups after four weeks, with no further significant changes at twelve weeks.

In MCI, reduced FC compared to healthy individuals is often considered a neurological feature of the condition (Owens et al., 2024). In particular, reductions in FC within the DMN may help differentiate MCI from Alzheimer’s disease, where decreases in FC between the DMN and other brain regions are more pronounced (Ibrahim et al., 2021; Wang et al., 2024). Conversely, hyperconnectivity in MCI has been interpreted as a compensatory remodeling of brain networks to offset functional decline (Grady et al., 2003; Liang et al., 2011; Pihlajamäki et al., 2009; Wang et al., 2006). Increased DMN activity has also been reported in MCI (Farràs-Permanyer et al., 2015), consistent with findings of elevated FC in early but not late MCI, when compensatory mechanisms may no longer be effective (Kang et al., 2021). In this context, therapeutic interventions could potentially reduce FC by diminishing compensatory hyperconnectivity. Similar interpretations have been proposed for reduced interhemispheric connectivity following repetitive transcranial magnetic stimulation (Ge et al., 2023). It is unlikely that the observed FC reductions in our study reflect a rapid deterioration of MCI over four weeks that then ceased, particularly as neuropsychological test results (reported elsewhere) indicated nonspecific symptom improvement rather than worsening.

We observed pronounced FC reductions in the anterior and posterior cingulate cortices, both key nodes of the DMN, which have consistently been implicated in MCI (Ibrahim et al., 2021; Pihlajamäki et al., 2009). Previous studies have reported both increases and decreases in FC in these regions in MCI (Cera et al., 2019; Wang et al., 2024; Yu et al., 2017; Zhan et al., 2018), making integration of findings challenging. Given that FC loss has been linked to increased risk of progression to Alzheimer’s disease (Bischkopf et al., 2002; Mitchell & Shiri-Feshki, 2009), and that both compensatory hyperconnectivity and disease-specific trajectories have been described, FC changes in MCI likely arise from multiple interacting mechanisms.

Although we did not detect vortioxetine-specific changes, several regions showing the strongest FC alterations across groups have previously been associated with MCI and with vortioxetine effects in depression. Temporal lobe atrophy and altered activation are well documented in MCI (for a review, see Pihlajamäki et al. (2009)). In our study, the transverse temporal gyrus showed the most pronounced FC reduction over time and with age. Reduced FC in ventral temporal association areas following acute vortioxetine administration, but not duloxetine, has been reported in rats (Pérez et al., 2018). Whether the temporal lobe FC reductions observed here involve early vortioxetine effects or reflect only nonspecific treatment-related changes remains unclear. Cross-species comparisons of temporal cortex homology remain debated (Xu et al., 2022; Xu et al., 2020), and the transverse temporal gyrus (Heschl’s gyrus), located in the primary auditory cortex, may not be affected by functional decline in the same way as medial temporal regions. FC of Heschl’s gyrus has been reported to increase in hearing loss (Fitzhugh et al., 2019; Fitzhugh & Pa, 2022) and to remain preserved in MCI compared to Alzheimer’s disease (Wang et al., 2020). Thus, the observed FC decrease in this region is unlikely to reflect hearing loss but may indicate age-related progression of MCI.

Among the few regions with increased FC, the cerebellum showed the strongest changes. Prior work in Parkinson’s disease linked increased cerebellar FC to compensatory mechanisms when patients developed additional cognitive impairment (Zhan et al., 2018). The cerebellum is now recognized as contributing to a wide range of functions beyond motor control, including cognition and neurodegenerative processes (Brissenden et al., 2018; Rudolph et al., 2023). With a functional organization resembling that of the cerebrum (Guell & Schmahmann, 2020), the cerebellum may provide compensatory support more effectively than cortical remodeling alone. Increases in cerebellar FC observed here, both over the first four weeks and with age, may reflect a distinct timeline of cerebellar involvement, potentially recruited when other compensatory mechanisms are insufficient or as part of therapy-related adaptations.

### Limitations

Our study did not include healthy controls or assessments prior to MCI onset, limiting interpretation of whether observed FC reductions represent reversal of compensatory hyperconnectivity. To minimize errors in region delineation due to atrophy, we used anatomically defined parcellations (DKT and Destrieux atlases). While such parcellations can reduce signal homogeneity in FC analyses (Gordon et al., 2016), they help avoid anatomical mismatches, and our agreement analysis suggests results were not specific to one atlas. Finally, although our fMRI acquisition parameters were optimized following Human Connectome Project recommendations (Smith et al., 2013), the high acceleration factor (multiband 8) may reduce signal-to-noise ratio in subcortical regions (Risk et al., 2021), potentially limiting detection of subcortical FC changes.

## Conclusion

We did not observe vortioxetine-specific FC changes in patients with MCI over twelve weeks of treatment. Instead, we found overall reductions in FC across both vortioxetine and placebo groups. These reductions may reflect reversal of compensatory hyperconnectivity, consistent with prior literature. The only previous study of vortioxetine in MCI reported positive findings, but lacked a control condition, raising the possibility of nonspecific effects. Vortioxetine-related FC changes described in depression may not generalize to MCI. Future studies should aim to distinguish compensatory hyperconnectivity from disease-related FC reductions by including healthy controls, longitudinal assessments before MCI onset, and comparisons between patients with MCI with and without comorbid depression. Such designs could clarify the contributions of disease and treatment to FC changes in MCI.

## Data availability

The data underlying the analyses in the present study will be made available upon reasonable request to the corresponding author after publication.

